# Key findings and recommendations from assessment of Cambodia’s national malaria surveillance system

**DOI:** 10.1101/2025.07.02.25330601

**Authors:** Siv Sovannaroth, Alexa Kugler, Pengby Ngor, Hannah Brindle, Bunmeng Chhun, Elijah Filip, Vanna Hem, Vichka Khy, Rafael Jairah Matoy, Vunsokserey Ou, Sany Ran, Pascal Ringwald, Kimleng Sok, Rattana Yoem, Zaixing Zhang, Rekol Huy

**Affiliations:** National Center for Parasitology, Entomology, and Malaria Control (CNM), Phnom Penh, Cambodia; Clinton Health Access Initiative, Phnom Penh, Cambodia; Clinton Health Access Initiative, Vientiane, Lao PDR; Mekong Malaria Elimination Programme, World Health Organization, Phnom Penh, Cambodia

## Abstract

Cambodia is approaching the final stages of malaria elimination, reporting only 1,384 cases in 2023 and aiming to eliminate all species of human malaria by 2025. Given the importance of surveillance in control and elimination efforts the National Center for Parasitology, Entomology, and Malaria Control (CNM) elected to assess its malaria surveillance system to identify strengths and gaps and make recommendations.

The WHO Malaria Surveillance System Assessment Toolkit, tailored to Cambodia’s context, was utilized to evaluate a total of 68 indicators across four primary objectives: Performance, Context and Infrastructure, Technical and Process, and Behavior. Data from 2021-2023 and activities which took place in 2023 under the 2021 edition of the Surveillance for Malaria Elimination Guidelines were utilized. Indicators were evaluated through a quantitative survey completed by subnational staff, a field data quality assessment (DQA) at the service delivery level, a national-level DQA, and a national-level desk review including interviews with CNM staff.

The assessment demonstrated strong performance of Cambodia’s surveillance system. Of the core indicators assessed, 82% of Performance (14/19), 77% of Context and Infrastructure (8/13), 79% of Technical and Process (4/7), and 100% of Behavior indicators (3/3) were met. Notable successes included the improved completeness and timeliness of reporting and case investigation in the previous three years. Key areas for improvement include revising case investigation and updating case classifications, enhancing data cleaning and verification processes to improve data quality, and upgrading the national malaria information system to include unique patient identifiers and enable interoperability with other national information systems.

Cambodia’s surveillance system is strong overall and CNM is committed to addressing identified areas of improvement. Lessons learned from this assessment informed revisions to the national Surveillance for Malaria Elimination Guidelines, implemented in May 2024. Additionally, the results will be used to guide the design of surveillance response-related objectives for Cambodia’s 2026-2035 National Strategic Plan.

## Introduction

Cambodia aims to eliminate all forms of malaria by 2025.[1] The country is making significant progress towards this goal, with only 1,384 cases reported in 2023, a 66% decrease from 2022 and 98% decline since 2018.[2] Recent years have focused on the elimination of *P. falciparum* due to its higher morbidity and mortality and the threat of artemisinin resistance in the region. As a result, in 2023 only 30 *P. falciparum* cases were recorded.

Malaria surveillance, defined by the World Health Organization as “the continuous and systematic collection, analysis and interpretation of malaria-related data, and the use of that data in the planning, implementation and evaluation of public health practice,” is a crucial component to achieve malaria elimination.[3] Routine monitoring and periodic evaluation of surveillance systems is essential to ensure their efficient and effective functioning. This is particularly important as countries move from malaria control to elimination and surveillance needs change.[4] The Malaria Surveillance Assessment Toolkit was developed by the WHO Global Malaria Program (GMP) with support from the Clinton Health Access Initiative (CHAI) and other partners to provide a ‘standardized but adaptable package of tools to assess surveillance systems across all transmission settings.’[5–7]

As Cambodia nears elimination, the National Malaria Control Program at the National Center for Parasitology, Entomology, and Malaria (CNM) elected to assess its malaria surveillance system to identify strengths and weaknesses within the system using the WHO Malaria Surveillance Assessment Toolkit, adapted to the Cambodia context. The aim of this assessment was to evaluate the performance of the national malaria surveillance system under CNM and the Malaria Information System (MIS). The results of this assessment were used to formulate a set of recommendations to strengthen the surveillance system and have informed revisions to the national malaria surveillance guidelines. Results will also be used to inform the next malaria National Strategic Plan (NSP) and contribute to national prevention of malaria re-establishment (PoR) guidelines currently under development.

## Methods

A total of 68 indicators (51 priority and 17 optional) from the WHO Malaria Surveillance Assessment Toolkit were selected for assessment under four primary objectives: Performance, Context and Infrastructure, Technical and Process, and Behavior.[6] Data from 2021-2023 and policies and interventions in place in 2023, under the 2021 edition of the Surveillance for Malaria Elimination Guidelines, were utilized.[8]

Indicators were evaluated through a quantitative survey completed by subnational staff, a field data quality assessment (DQA) undertaken at the service delivery level and a national-level DQA, and a national-level desk review including interviews with CNM staff. The desk review tool provides criteria for categorizing each priority indicator result as met, partially met, or not met, which is translated into a score for each objective and sub-objective in the Elimination Scorecard.

### Quantitative survey

A survey was administered to a total of three Provincial Malaria Supervisors (PMS), four Operational District Malaria Supervisors (ODMS), staff at nine health facilities (HF) and 88 VMWs in three provinces: Battambang (low malaria burden), Kampong Speu (moderate burden), and Mondulkiri (high burden).

### Service-delivery level data quality assessment

A service delivery-level DQA of core malaria variables was conducted at the health center (n=9) and community levels (n=88) in the same provinces as the quantitative survey. Data recorded in registers were compared to data reported to the MIS, for the most recent ten cases reported by each point of care. Data for 2023 were analyzed for completeness and concordance of core variables. Concordance was calculated by the exact match of all corresponding fields per case in the MIS line listed data and the paper registers for a total of 14 core variables. Given the potential for spelling errors, concordance was also calculated excluding location-specific fields. Core variables were adapted from those specified in the WHO Surveillance Assessment Toolkit and included: patient residence, diagnosis date, diagnosis method, species, treatment date, treatment, notification date, recent travel in country, in-country travel location, recent travel outside country, outside country travel location, classification, case investigation date, case investigation GPS.

### Desk review

A comprehensive desk review of national program documents, reports, presentations, and records was conducted at the national level. This was supported by qualitative interviews and consultation with individuals from the National Program representing various CNM units and focal points.

### Desk-level data quality assessment

A desk-level DQA was conducted for data timeliness, completeness, and consistency at the national, provincial, district, and health facility levels. This DQA was completed using country-wide case line-listed data from endemic operational districts between 2021-2023.

## Results

According to the Elimination Scorecard, Cambodia met all Behavior indicators (3/3), 82% of Performance indicators (14/19), 79% of Technical and Process indicators (4/7), and 77% of Context and Infrastructure indicators (8/13). Not all indicators selected for this assessment are included in the WHO scorecard; the full results by indicator can be found in Table 1. Below we discuss the key findings and recommendations (Table 2).

**Table 1.**
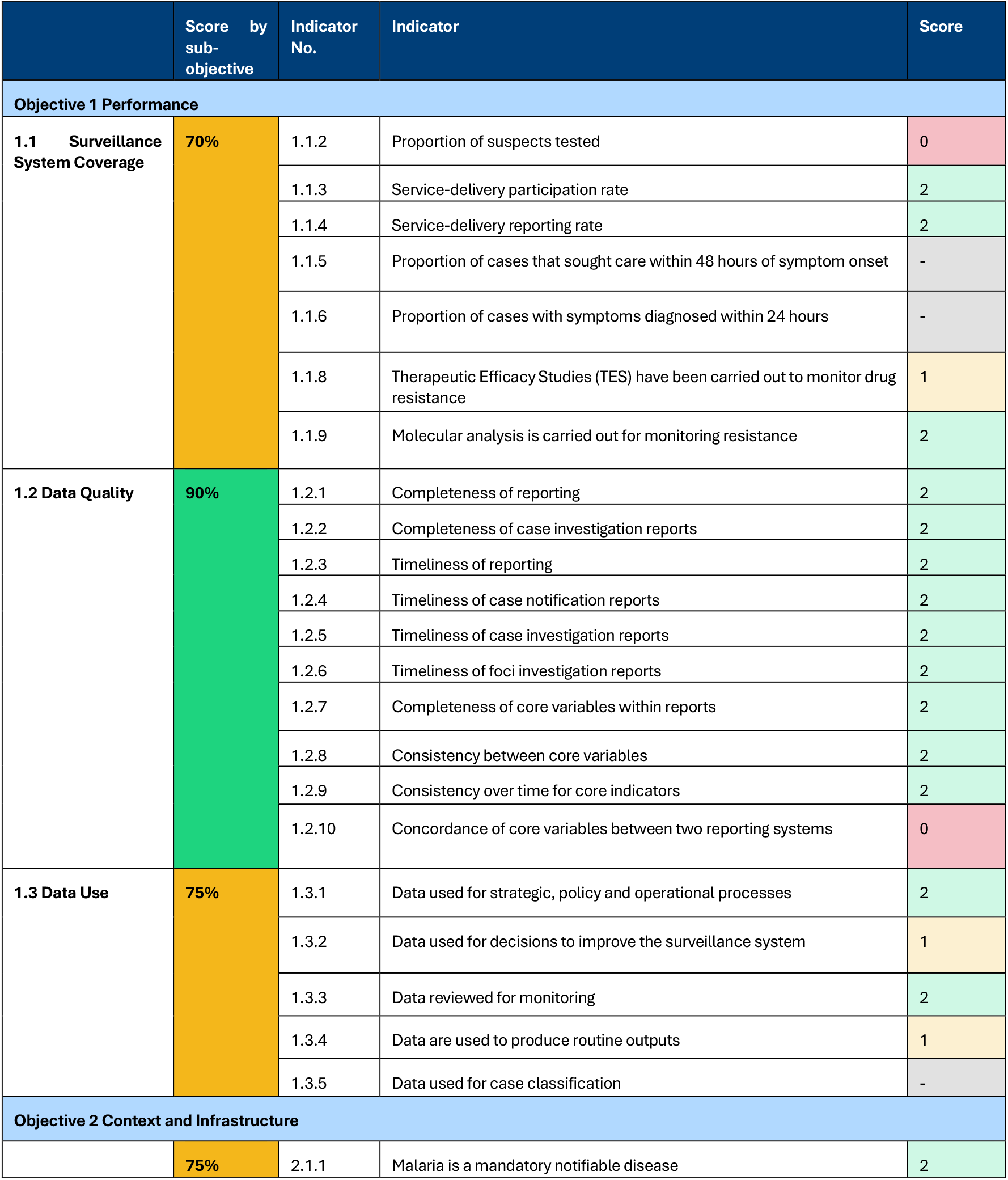
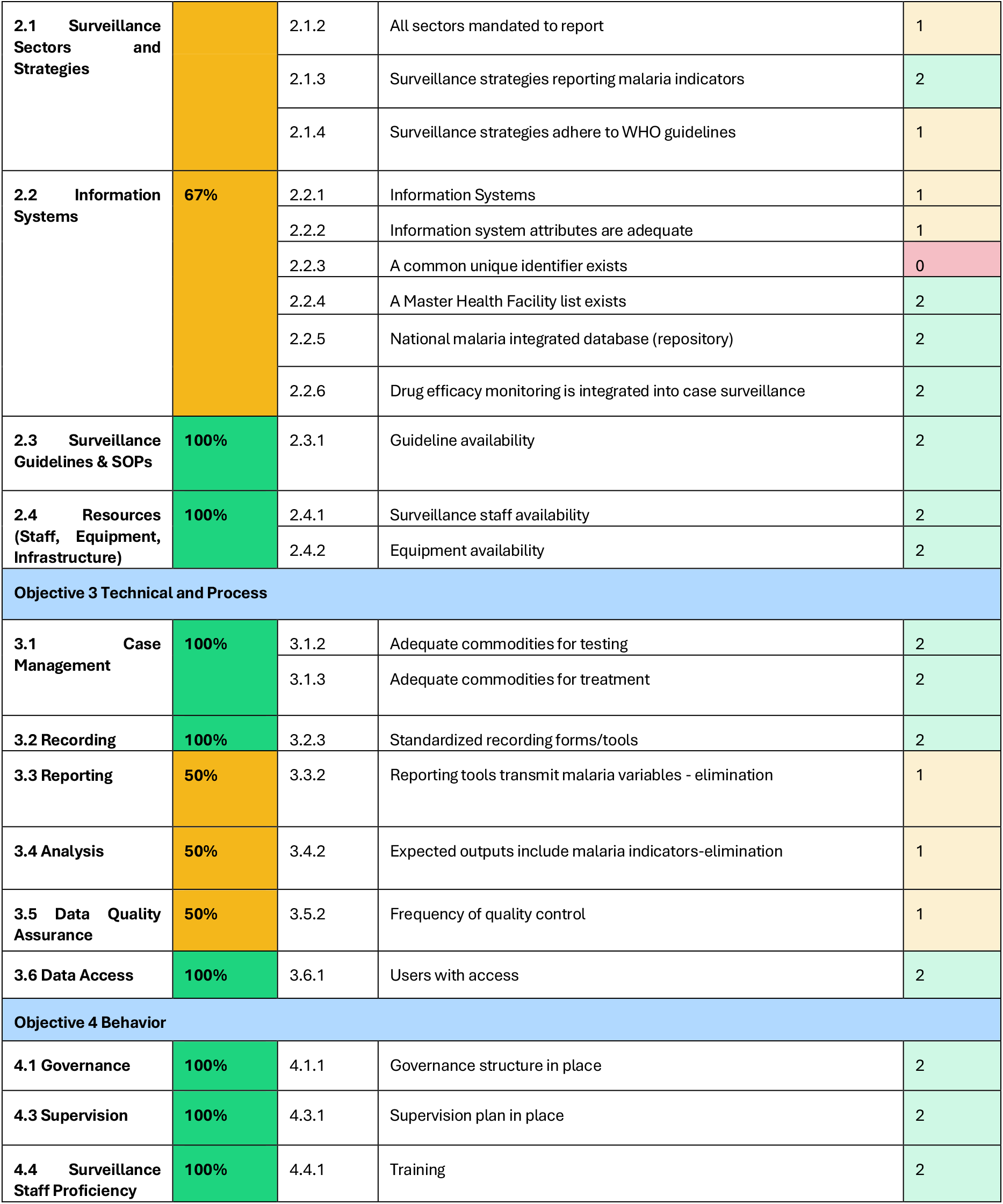
Summary scorecard of key indicators. A score of 0 = Not met, 1 = Partially met, 2 = Met, and – denotes that the indicator was not assessed.

|  | Score by sub-objective | Indicator No. | Indicator | Score |
| --- | --- | --- | --- | --- |
| Objective 1 Performance |  |  |  |  |
| 1.1 Surveillance System Coverage | 70% | 1.1.2 | Proportion of suspects tested | 0 |
|  |  | 1.1.3 | Service-delivery participation rate | 2 |
|  |  | 1.1.4 | Service-delivery reporting rate | 2 |
|  |  | 1.1.5 | Proportion of cases that sought care within 48 hours of symptom onset | - |
|  |  | 1.1.6 | Proportion of cases with symptoms diagnosed within 24 hours | - |
|  |  | 1.1.8 | Therapeutic Efficacy Studies (TES) have been carried out to monitor drug resistance | 1 |
|  |  | 1.1.9 | Molecular analysis is carried out for monitoring resistance | 2 |
| 1.2 Data Quality | 90% | 1.2.1 | Completeness of reporting | 2 |
|  |  | 1.2.2 | Completeness of case investigation reports | 2 |
|  |  | 1.2.3 | Timeliness of reporting | 2 |
|  |  | 1.2.4 | Timeliness of case notification reports | 2 |
|  |  | 1.2.5 | Timeliness of case investigation reports | 2 |
|  |  | 1.2.6 | Timeliness of foci investigation reports | 2 |
|  |  | 1.2.7 | Completeness of core variables within reports | 2 |
|  |  | 1.2.8 | Consistency between core variables | 2 |
|  |  | 1.2.9 | Consistency over time for core indicators | 2 |
|  |  | 1.2.10 | Concordance of core variables between two reporting systems | 0 |
| 1.3 Data Use | 75% | 1.3.1 | Data used for strategic, policy and operational processes | 2 |
|  |  | 1.3.2 | Data used for decisions to improve the surveillance system | 1 |
|  |  | 1.3.3 | Data reviewed for monitoring | 2 |
|  |  | 1.3.4 | Data are used to produce routine outputs | 1 |
|  |  | 1.3.5 | Data used for case classification | - |
| Objective 2 Context and Infrastructure |  |  |  |  |
|  | 75% | 2.1.1 | Malaria is a mandatory notifiable disease | 2 |
| 2.1 Surveillance Sectors and Strategies |  | 2.1.2 | All sectors mandated to report | 1 |
|  |  | 2.1.3 | Surveillance strategies reporting malaria indicators | 2 |
|  |  | 2.1.4 | Surveillance strategies adhere to WHO guidelines | 1 |
| 2.2 Information Systems | 67% | 2.2.1 | Information Systems | 1 |
|  |  | 2.2.2 | Information system attributes are adequate | 1 |
|  |  | 2.2.3 | A common unique identifier exists | 0 |
|  |  | 2.2.4 | A Master Health Facility list exists | 2 |
|  |  | 2.2.5 | National malaria integrated database (repository) | 2 |
|  |  | 2.2.6 | Drug efficacy monitoring is integrated into case surveillance | 2 |
| 2.3 Surveillance Guidelines & SOPs | 100% | 2.3.1 | Guideline availability | 2 |
| 2.4 Resources (Staff, Equipment, Infrastructure) | 100% | 2.4.1 | Surveillance staff availability | 2 |
|  |  | 2.4.2 | Equipment availability | 2 |
| Objective 3 Technical and Process |  |  |  |  |
| 3.1 Case Management | 100% | 3.1.2 | Adequate commodities for testing | 2 |
|  |  | 3.1.3 | Adequate commodities for treatment | 2 |
| 3.2 Recording | 100% | 3.2.3 | Standardized recording forms/tools | 2 |
| 3.3 Reporting | 50% | 3.3.2 | Reporting tools transmit malaria variables - elimination | 1 |
| 3.4 Analysis | 50% | 3.4.2 | Expected outputs include malaria indicators-elimination | 1 |
| 3.5 Data Quality Assurance | 50% | 3.5.2 | Frequency of quality control | 1 |
| 3.6 Data Access | 100% | 3.6.1 | Users with access | 2 |
| Objective 4 Behavior |  |  |  |  |
| 4.1 Governance | 100% | 4.1.1 | Governance structure in place | 2 |
| 4.3 Supervision | 100% | 4.3.1 | Supervision plan in place | 2 |
| 4.4 Surveillance Staff Proficiency | 100% | 4.4.1 | Training | 2 |

**Table 2.** Recommendations to address identified gaps and strengthen national malaria surveillance system.

| Recommendation | Associated indicators |
| --- | --- |
| **Guidelines and data collection** |  |
| Collect the date/time of symptom onset for malaria patients | 1.1.5, 3.2.2 |
| Collect the date/time of presentation to care | 1.1.5, 1.1.6 |
| **Collect a detailed travel history data for all cases, while considering the likely period of infection** | 1.3.5, 3.2.2 |
| **Update case classifications to include indigenous, introduced, induced, and recrudescent** | 2.4.1, 2.3.2, 3.3.2, 3.4.2 |
| Disaggregate cases by inpatient and outpatient | 3.2.2, 3.3.2, 3.4.2 |
| Calculate and report additional WHO-recommended indicators for which CNM already collects the necessary data, if helpful to the national program (e.g. proportion of population living in active foci) | 3.4.2 |
| **Include guidance on data storage, management in the POR and SNV guidelines, as well as the next National Strategic Plan** | 2.3.2 |
| **Require the military, police, and non-endemic health centers to report real-time, case-based data** | 2.1.2 |
| **Data quality, review, and use** |  |
| **Include additional data verification (completeness of core variables, concordance between registers and MIS of additional variables) in routine supervision visits** | 1.2.12, 3.5.1 |
| **Incorporate additional data verification and identification of the root cause of errors and data quality issues in the monthly/quarterly VMW meetings** | 1.2.12 |
| Resume the surveillance partner review meetings | 1.3.3 |
| **Expand routine data cleaning activities at the national level from verification of case species to include routinely checking for missing values, duplicate entries, outliers, unexpected values, etc** | 3.5.1 |
| **Expand monitoring of data quality indicators to include completeness/missingness of specific core variables, concordance between register and MIS data, monitoring for outliers, etc** | 3.5.1 |
| Update the existing MIS guide to include information on data security | 2.3.2 |
| **Increase the collaboration between MIS and HMIS to ensure completeness of reporting and capture of all malaria cases across endemic and non-endemic operational districts** | 1.2.10 |
| **Subnational staff capacity building** |  |
| Include subnational monitoring of surveillance data in the context of progress towards HC-level elimination targets and SNV | 1.3.1, 1.3.2 |
| Introduce supervision e-checklist at the subnational level, including scoring, and utilize score to inform feedback and supervision schedule | 1.3.2 |
| Incorporate in the next MIS training to subnational levels (PHD, OD, HC, and VMW): | 1.3.4, 3.3.4, 4.4.1, 4.4.3 |
| • Refresher training on use of MIS dashboards<br>• Basic data cleaning, analysis, and use for decision making, including calculation of standard malaria indicators and preparation of data visuals for reporting<br>• Refresher on case-based data reporting, reporting of zero cases |  |
| Ensure all VMW have current job aids | 4.4.2 |
| **Malaria Information System (MIS) strengthening** |  |
| Implement a helpdesk and ticket system for the MIS in alignment with ITIL (Information Technology Infrastructure Library) framework or other industry standards; for submitting and triaging user issues, suggestions, and requests, and tracking the decision-making and implementation of changes | 2.2.2 |
| Establish a routine maintenance plan (with costing) and SOPs for the MIS including daily/weekly/monthly/annual tasks, backup and recovery, monitoring performance, documentation and change management, incident response plan, etc in alignment with international standards and methodologies (eg ITIL, ISO/IEC, Agile or Scrum frameworks) | 2.2.2 |
| Implement a unique patient ID system to enable identification of duplicate records and relapsing or re-infected patients | 2.2.3, 3.2.2, 3.5.3 |

### Description of the surveillance system

Malaria is a mandatory notifiable disease in Cambodia; all public providers must report malaria cases, and the private sector must refer suspect cases to the public sector. Public facilities in the country’s 55 malaria-endemic operational districts (OD) report case-based data to the CNM Malaria Information System (MIS) in real-time, while the remaining 48 ‘non-endemic’ OD report aggregate monthly data to the Health Management Information System (HMIS) under the department of health information and planning (DPHI). Data from endemic areas includes line-listed case investigation and classification, focus investigation and classification, and other malaria control intervention data. Data in the HMIS are aggregated according to inpatients/outpatients, severe/simple malaria, sex, and age group; and cases are not investigated nor classified.

### Malaria Information System

The MIS, first launched in 2010 as a desktop application and transitioned in 2018 to a real-time reporting system via mobile applications, was cited by CNM staff as crucial to improve data quality, availability, and use for decision making. However, currently, the system lacks standardized routine maintenance. Due to this 22% (n=2) of surveyed health facilities reported an information system failure in the two weeks prior preventing the collection or reporting of data.

### Data quality

Cambodia employs a 1-3-7 approach where cases must be notified, investigated, and classified within 24 hours, reactive case detection conducted within 3 days, and focus investigation completed within 7 days. Completion of case investigation increased from 90.3% of confirmed cases investigated in 2021 to 99.6% investigated in 2023. In 2023, 87% of cases were notified within 24 hours, increasing from 37% in 2021 to 63% in 2022. Timeliness of focus investigations within 7 days increased from 29% in 2021 (12/41) to 100% in 2023 (4/4).

While data validation rules are built into the MIS system and an audit trail is in place, data cleaning is limited to validation of the malaria species for each case. Monitoring of data quality indicators in the MIS consists of completion (submission) of monthly reports and timeliness of 1-3-7 activities. Of the subnational respondents, 57% reported systematically undertaking any data quality indicator monitoring, primarily undertaken as part of routine supervision which was reported by all provinces and ODs surveyed and performed monthly by 13/16 (81%). In practice, data quality monitoring during supervision visits consists of validating the number of cases and tests reported to the MIS against register data.

For 55/59 (93%) of cases from 2023 included in the service delivery level DQA, all 14 core variables were complete in the paper registers. However, there was discordance between the entry of core variables in the paper-based registers and the MIS. Among 56 cases reviewed for concordance between the paper registries and MIS, core variables were fully concordant in 7 cases (13%). The in-country travel location was the most highly discordant variable (discordant in 25 (47%) cases), followed by treatment provided (discordant in 11 cases (21%)). Following exclusion of location-specific variables, 16 (29%) cases were fully concordant.

### Data use

The most common use of data was to develop subnational operational plans (100%, n=7) followed by stratification for targeting and prioritizing interventions (86%, n=6). Data were less frequently used for the distribution of commodities or to advocate for a policy or program (43% (n=3) and 29% (n=2), respectively). Data were also used to improve the surveillance system including feedback and supervision processes, data quality and to initiate trainings/capacity development (57% of responses (n=4) in each case) and to produce routine outputs, most commonly monthly bulletins at the district and health facility level (83.3% and 111.1% of bulletins produced out of those expected during 2023, respectively).

## Discussion

The WHO malaria surveillance assessment toolkit provided an important opportunity to review the surveillance system in Cambodia, the first time it has performed a standardized assessment on this scale. Prior examples include a self-administered survey of the sources of malaria surveillance data conducted in 2016.[9] The assessment demonstrated that performance was strong overall, however some areas for improvement were identified, from which recommendations to support the achievement of malaria elimination were drafted by the national program (Table 2).

Given routine supervision visits were identified as a strength of the surveillance system, they should be utilized for additional data quality checks, especially concordance of additional key variables. Additionally, the monthly and quarterly VMW meetings can be used for data verification and identification of root cause of discrepancies between the MIS and registers. To further strengthen the MIS, a routine maintenance plan in alignment with international standards in information systems should be developed and implemented to improve system stability and security. In addition, a mechanism for submission of user feedback, errors, and suggestions should be developed. At the national level, routine data cleaning activities should be expanded to include checking for missing values, duplicate entries, outliers and unexpected values, and other routine data cleaning procedures. Resumption of surveillance partners meetings could serve as an additional mechanism for monitoring the improvement of data quality.

Although this was a comprehensive assessment of the MIS across multiple levels, the HMIS was not included in the scope of this assessment which was a limitation. Results should be interpreted within the context of the endemic operational districts. Should a future assessment be conducted, we would recommend including HMIS, with a particular focus of the interoperability between the two systems to ensure all cases are captured in a timely manner, investigated and classified.

Furthermore, the private sector was not included in this assessment. Although private providers cannot diagnose or treat malaria, they are responsible for referring suspect malaria cases to the public sector. Prior to the close of the Public-Private Mix (PPM) program in 2018, the private sector accounted for nearly 40% of annual cases detected.[10] And, according to a 2013 survey, while nearly 90% of people sought treatment for fever, the majority (58%) sought care from the private sector (58%).[11] The most recent data, from a 2020 survey, showed that care-seeking behavior may have shifted. Respondents’ first choice for seeking testing for illness they suspect to be malaria is at a health center or hospital (57.2%), VMW/MMW (23.7%), or private provider (13%).[12]

Further evaluation of the referral system between private and public sector is warranted to ensure that all suspected malaria cases receive a parasitological diagnosis.

We would recommend additional investigation of certain components of the assessment. Discordance between the MIS and paper-based registries was highlighted as a considerable issue and may have affected assessment results based on routinely collected data. Although further investigation into the cause of these discrepancies is warranted to ensure that data entry errors during DQA have not contributed to the magnitude of discordance, during qualitative interviews with national program staff, it was hypothesized that some discrepancies could be due to variation in fields or field options between paper registers and the MIS reporting app, creating confusion at the point of care level. Routine supervision visits and routine VMW meetings can therefore be utilized to delve into the specific causes behind any discrepancies and identify solutions to improve concordance in the future.

In conclusion, in 2023 Cambodia undertook its first comprehensive evaluation of the malaria surveillance system administered by CNM. The results were largely positive, highlighting the strength and progress of malaria surveillance in Cambodia. Two key gaps were identified related to system comprehensiveness and data quality, and plans have been initiated to address these. Results and recommendations are informing changes to Cambodia’s surveillance system and will help Cambodia prepare for successful malaria-free certification.

## Disclaimer

PR is staff member of WHO. The author alone is responsible for the views expressed in this publication, which do not necessarily represent the decisions, policies, or views of WHO.

## Data Availability

National malaria surveillance data are available at https://mis.cnm.gov.kh/. Additional data available upon reasonable request to the National Center for Parasitology, Entomology and Malaria Control (CNM), Phnom Penh, Cambodia.

## References

1. National Center for Parasitology, Entomology, and Malaria Control (CNM), Cambodia Ministry of Health. Cambodia Malaria Elimination Action Framework 2021-2025 [Internet]. National Center for Parasitology, Entomology, and Malaria Control (CNM); 2021 [cited 2024 Aug 7]. Available from: https://mis.cnm.gov.kh/media/documents/6518B6BBB20D6A903AC4387FB7EA5417.pdf

2. National Center for Parasitology, Entomology, and Malaria Control (CNM). Malaria Information System (MIS). 2024 [cited 2024 Aug 7]. Malaria Dashboard V2. Available from: https://mis.cnm.gov.kh/Dashboard

3. World Health Organization. World Health Organization. 2024 [cited 2024 Sep 19]. Global Malaria Programme. Available from: https://www.who.int/teams/global-malaria-programme/surveillance

4. World Health Organization Global Malaria Programme. A framework for malaria elimination [Internet]. World Health Organization; 2017 Feb [cited 2024 Nov 1]. Available from: https://www.who.int/publications/i/item/9789241511988

5. Global Malaria Programme (GMP). Malaria surveillance assessment toolkit implementation reference guide [Internet]. World Health Organization; 2022 [cited 2024 Sep 8]. Available from: : https://www.who.int/publications/i/item/9789240055278

6. World Health Organization. Malaria Surveillance Assessment Toolkit [Internet]. [cited 2024 Aug 9]. Available from: https://malsurtoolkit.who.int/

7. World Health Organization. Surveillance system assessments [Internet]. [cited 2024 Sep 25]. Available from: https://www.who.int/teams/global-malaria-programme/surveillance/surveillance-system-assessments

8. National Center for Parasitology, Entomology, and Malaria Control (CNM), Cambodia Ministry of Health. Cambodia Surveillance for Malaria Elimination, 2021 Edition [Internet]. National Center for Parasitology, Entomology, and Malaria Control (CNM); 2021 [cited 2024 Sep 4]. Available from: https://cnm.gov.kh/index.php?&action=ID203

9. Mercado CEG, Ekapirat N, Dondorp AM, Maude RJ. An assessment of national surveillance systems for malaria elimination in the Asia Pacific. Malar J. 2017 Mar 21;16(1):127.

10. National Center for Parasitology, Entomology, and Malaria Control (CNM), Cambodia Ministry of Health. Cambodia Malaria Elimination Action Framework 2016-2020 [Internet]. National Center for Parasitology, Entomology, and Malaria Control (CNM); 2016 [cited 2024 Sep 14]. Available from: https://cnm.gov.kh/index.php?&action=ID203

11. National Center for Parasitology, Entomology, and Malaria Control (CNM), Malaria Consortium, UN Office for Project Services. Cambodia Malaria Survey 2013 [Internet]. 2013. Available from: https://www.malariaconsortium.org/media-download-273 file/201602290241/-/report_cambodiamalariasurvey_2013.pdf

12. National Center for Parasitology, Entomology, and Malaria Control (CNM), Center for Health and Social Development (HSD). Cambodia Malaria Survey 2020. 2020.

